# Time-Surrogate Variables Enhance the Association Between Cardiotocographic Features and Intrapartum Hypoxic-Ischemic Encephalopathy

**DOI:** 10.1101/2025.11.16.25340361

**Authors:** Johann Vargas-Calixto, Michael W. Kuzniewicz, Marie-Coralie Cornet, Yvonne W. Wu, Aditi Lahiri, Lawrence Gerstley, John Parker, Philip A. Warrick, Robert E. Kearney

## Abstract

**Background:** Interruptions to the flow of oxygenated blood to the fetal brain during labor can lead to hypoxic-ischemic encephalopathy (HIE). Timely preventive interventions for suspected hypoxemia are crucial to avoid progression to neurological injury. Prior research has used features from fetal heart rate (FHR) and uterine activity (UA) signals to predict adverse fetal outcomes. However, these systems focused only on the end of labor, when preventive measures are unlikely effective. Previously, we demonstrated that accounting for proximity to birth improved the association between FHR and UA features and the outcome group. Although proximity to birth cannot be known prospectively, other time-surrogate variables (TSVs) may be as useful.

**Methods:** We analyzed intrapartum data from 152,761 vaginal births, comprising 150,813 with healthy outcomes, 1,793 with perinatal acidosis, and 155 with confirmed HIE. Classical and novel FHR features were extracted across labor durations of up to 72 hours. This dataset represents the largest cohort of intrapartum FHR and UA signals to date, both in participant count and signal length. We evaluated four alternative TSVs to evaluate their ability to enhance the association of CTG features and the development of HIE. To do so, we applied information-theoretic methods to quantify their contribution to the association of fetal outcomes with FHR and UA features.

**Findings:** Our results show that the cumulative contraction duration and the time from labor onset (TLO) were the most effective TSVs for strengthening the association FHR, UA, and fetal outcome. However, it is more practical to track TLO in clinical settings than a continuous contraction monitoring.

**Conclusion:** TLO is the most suitable TSV for prospective intrapartum CTG evaluation. Incorporating it may substantially enhance the performance of automated systems for early detection of intrapartum HIE.

**Author summary:** Our study addressed a significant challenge in the development of classifiers to detect fetuses at high risk of developing hypoxic-ischemic encephalopathy (HIE) during labor. Most previous classifiers have concentrated on the end of labor, overlooking the dynamic evolution of FHR and UA patterns. This limits their clinical utility, as preventive interventions late in labor are unlikely to avert adverse fetal outcomes. To address this gap, we evaluated several TSVs that capture the progression of FHR and UA patterns and could support prospective, clinically usable intrapartum classifiers. Analyzing data from over 150,000 births, we found that time since labor onset was the most informative and practical TSV for enhancing the associations between FHR and UA features and the development of HIE during labor. Incorporating this variable would enable classifiers to learn the temporal dynamics of FHR and UA features, potentially improving early identification of fetuses at elevated risk for HIE and supporting more timely, effective interventions.

## Introduction

During labor, uterine contractions intermittently compress maternal blood vessels that deliver oxygen and nutrients to the placenta and fetus [1]. These episodes result in transient reductions in oxygenated blood flow to the fetus. While the fetus can typically tolerate such events through a range of physiological and anatomical adaptations [2], these compensatory mechanisms may be overwhelmed when episodes are prolonged, recurrent, or severe. In such cases, the fetus may develop progressive hypotension and cerebral ischemia, and ultimately neonatal hypoxic-ischemic encephalopathy (HIE). Although HIE is relatively rare, it carries significant consequences, as approximately half of infants with moderate to severe HIE either die or suffer permanent neurological impairments [3, 4].

Detecting fetal deterioration leading to HIE during labor is difficult. There is no method for continuous assessment of fetal oxygenation and cerebral perfusion. However, fetal heart rate (FHR) is available and is modified by internal biosensors that react to oxygen levels and hypotension. Cardiotocography (CTG) refers to the clinical monitoring of FHR and maternal uterine activity (UA). The goal of CTG is to assess fetal tolerance to labor by identifying FHR patterns that indicate significant fetal hypoxemia, so that timely interventions, such as emergency Caesarean deliveries, can prevent progression to organ injury and neonatal HIE.

Clinical interpretation of FHR relies on visual assessment of three primary patterns: (1) baseline FHR and its variability, (2) accelerations, and (3) decelerations, which are further classified based on their morphology and timing relative to uterine contractions [5]. Accelerations, along with normal baseline levels and variability, are typically associated with favorable outcomes. Decelerations are a common physiological response to contractions; however, certain subtypes may signal significant hypoxia, altered fetal blood pressure, or myocardial compromise [6]. Despite these associations, over 80% of CTG tracings display FHR patterns that are neither clearly reassuring nor definitively indicative of hypoxemia [7]. As a result, current CTG interpretation has low sensitivity for identifying fetuses at increased risk of hypoxia-related complications such as HIE.

Computerized methods for CTG interpretation have yet to achieve clinical utility. One major limitation is the lack of accessible, large-scale databases containing sufficient cases of HIE to train robust and generalizable classifiers. Additionally, most current approaches focus on CTG data near delivery or at fixed time points, such as labor onset or conclusion, using time-invariant models that fail to account for the nonstationary nature of FHR and UA patterns.

In a recent study, we demonstrated that incorporating the proximity to birth, quantified as the time to delivery (TTD), significantly improved the association between CTG features and labor outcomes compared to approaches that excluded temporal context [8]. This finding suggests that time-aware classifiers could enhance the detection of intrapartum HIE.

However, the clinical use of TTD presents practical limitations. TTD is only known after birth and cannot be applied prospectively in real-time clinical settings. Moreover, its interpretation varies by delivery mode: in vaginal births, TTD reflects the natural progression of labor, while in Caesarean deliveries, birth timing is determined by clinical decision-making and often occurs earlier than it would have naturally.

Given the limitations of TTD, it remains unclear how to prospectively capture the temporal evolution of FHR and UA during labor. This raises the question: are there alternative time surrogate variables (TSVs) that could be used in real time and overcome the constraints of TTD? An ideal TSV would (1) correlate with CTG features, (2) enhance their association with fetal outcomes, (3) remain consistent across delivery methods, and (4) be applicable in real-time clinical settings.

In this study, we evaluated four candidate TSVs: A) Time from labor onset (TLO): measures the duration the fetus has been exposed to labor, B) Cumulative deceleration time (CDT): quantifies the total duration of hypoxic episodes reflected in FHR decelerations, C) Cumulative contraction time (CCT): captures the total time of uterine contractions, which may contribute to fetal hypoxia, and D) Contraction rate (CR): reflects the frequency of potentially hypoxic events over time. To assess the dependency of FHR and UA on these TSVs, we extracted 40 CTG features previously shown to be associated with HIE outcomes [8]: 14 from baseline FHR segments, 3 from acceleration segments, 20 from deceleration segments, and 3 from the UA signal. We then computed the normalized mutual information (NMI) between each TSV, the CTG features, and labor outcomes. Bootstrapping was used to estimate 95% confidence intervals for NMI values. Finally, we compared the performance of the four candidate TSVs against TTD and a stationary approach that excluded time information.

## Materials and methods

### Study population

This retrospective study analyzed clinical and CTG data from 15 Kaiser Permanente Northern California hospitals, collected between 2011 and 2019. Inclusion criteria were limited to singleton live births at term and late preterm gestation (35–40 weeks) without major congenital or chromosomal anomalies. The final dataset comprised 297,280 births. We examined all available intrapartum CTG recordings, which extended up to 72 hours prior to delivery. The Research Ethics Boards (REB) of Kaiser Permanente (approval numbers 1470283-5 and 1276201-29) and McGill University (approval numbers A04-M27-20A (20-04-025) and A04-M29-20B (20-04-056)) reviewed the study and determined that it involved secondary research of data for which consent was not required.

Fig 1 illustrates the breakdown of infants included in the database and those selected for analysis. We reviewed all available CTG data to identify the onset of labor [9]. Cases in which labor onset could not be determined were excluded, along with CTG segments recorded prior to labor onset. Additionally, we excluded fetuses delivered via Caesarean section to focus exclusively on the natural progression and culmination of labor for TSV comparisons.

**Fig 1.**
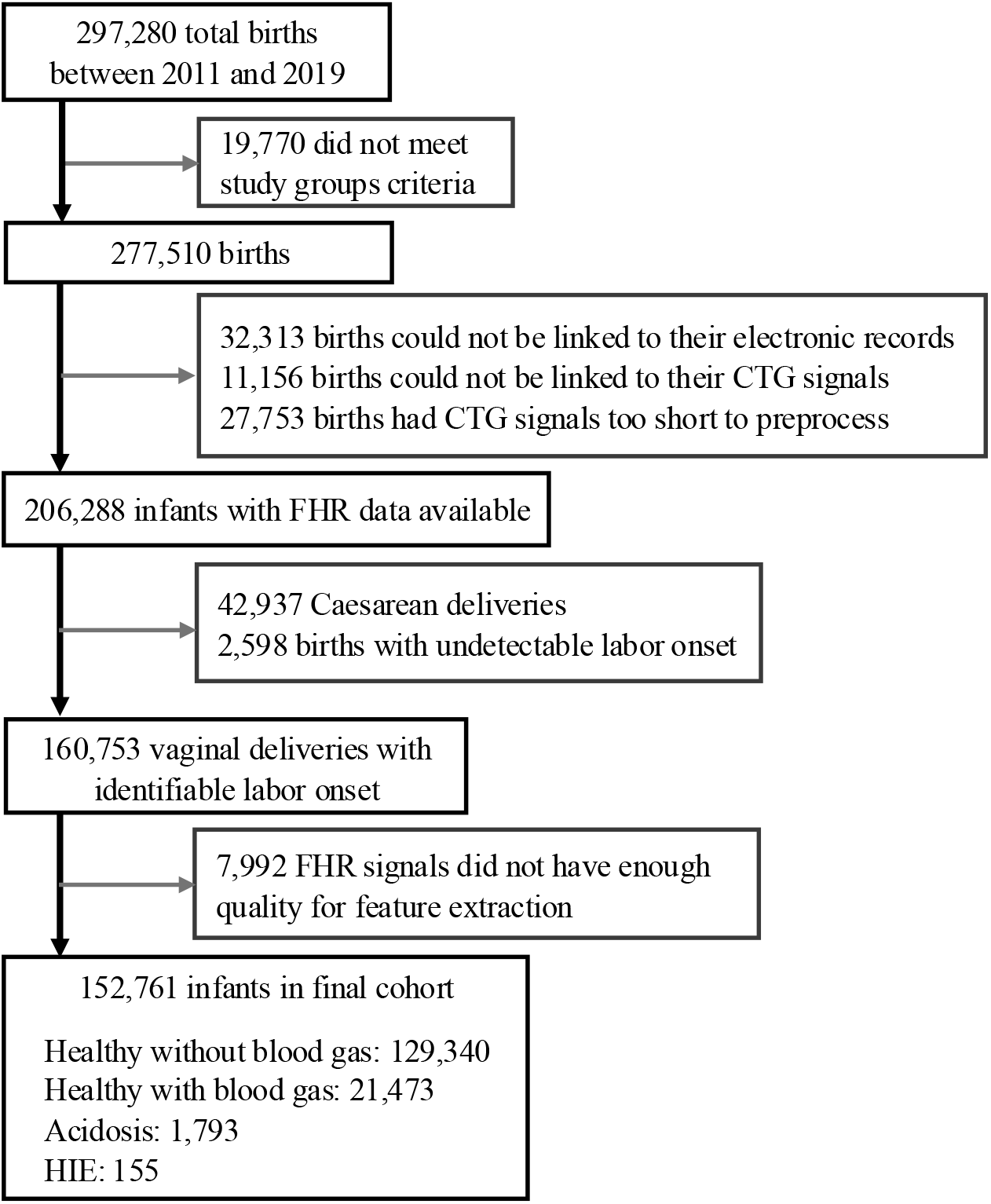
Inclusion and exclusion criteria of the infants included in the study.

The study population was first divided into two groups: those for which blood gases (BG) were collected, and those for which it was not, the majority. Blood gas collection is not the standard for all deliveries in Kaiser Permanente. Clinicians only collect umbilical or neonatal BG when they suspect fetal compromise during labor. The BG results were used to define whether the newborns suffered fetal hypoxemia that progressed to acidosis, a biomarker of HIE, during the intrapartum period. We included those infants that had a healthy outcome without BG data available (*n*=129,340). In this study, the pathological groups were defined based on a strict criteria in addition to the BG results. Infants who were encephalopathic, experienced seizures, or died but did not have blood gas data were excluded from the study, as a hypoxic etiology could not be definitively confirmed without evidence of acidosis on the blood gas. This subgroup did not meet the criteria to be included in the other groups. We distinguished three mutually exclusive outcome groups from infants with BG data assessments [9, 10]:

1. <u>The intrapartum HIE group (*n*=155)</u> presented acidosis and neonatal encephalopathy at birth. Acidosis was defined by a cord gas pH *<* 7.0 or base deficit (BD) ≥ 10 mmol/L, or neonatal gas BD ≥ 10 mmol/L in the first 2 hours after birth. Neonatal encephalopathy was determined by an abnormal neurological examination during the first six hours of life, associated with one of the following:
  - neurological abnormalities lasting beyond six hours of life,
  - neonatal seizures during the first day of life, or
  - the newborn requiring therapeutic hypothermia. All infants suspected of HIE underwent a careful chart review by a panel of clinicians to confirm the diagnosis [10].
2. <u>The acidosis group (*n*=1,793)</u> had acidosis at birth but did not meet the stringent criteria for HIE. Some were cooled or had seizures but no encephalopathy in the first six hours after birth.
3. <u>The healthy group (*n*=21,473)</u> had normal blood gases defined by a cord-gas pH ≥ 7.0 and BD *<* 10 mmol/L, or neonatal-gas BD *<* 10 mmol/L. In addition, other neonatal markers were reassuring:
  - the Apgar score at five minutes after birth was ≥ 7,
  - they were not admitted to the neonatal intensive care unit,
  - they were discharged home,
  - no death or hospital transfers in the neonatal period,
  - no evidence of NE,
  - they did not require intubation or chest compression during newborn resuscitation,
  - they did not suffer from seizures or receive medication for seizures, and
  - they did not require therapeutic hypothermia.

We treated the healthy without blood gas and the healthy with blood gas as distinct groups. This was because healthy infants with blood gas data may have had worrisome CTG patterns, which prompted the clinical assessment of their blood gases. Thus, differentiating this group from the pathological cases may be more difficult than differentiating those with no blood gas data.

### CTG signal processing

#### Preprocessing

The FHR and UA signals were sampled at 4 Hz. We preprocessed the FHR signal using PeriCALM Patterns, a software system developed by PeriGen Inc., that marked segments with high noise levels. These noisy segments were left as gaps in the signals. Gaps shorter than 60 samples (15 s) were linearly interpolated. Then, PeriCALM Patterns filtered the FHR using low-pass, high-pass, median filters, and a Karhunen-Loève filter to reduce the noise in the signals [11, 12].

#### Identification of CTG events and segmentation

After preprocessing, PeriCALM Patterns used a long-short-term memory network classifier to identify and label the CTG events that are the focus of clinical assessment [11, 12]. These included:

- <u>FHR baselines:</u> relatively flat segments of the FHR, typically in the range of 110 to 160 bpm and peak-to-peak variability between 5 and 15 bpm.
- <u>FHR accelerations:</u> transitory increases in FHR by more than 15 bpm from baseline, which last longer than 15 seconds before returning to baseline level.
- <u>FHR decelerations:</u> transitory decreases in the FHR by more than 15 bpm from the baseline, which last for more than 15 seconds before returning to the baseline level.
- <u>Uterine contractions:</u> gradual increases of energy in the UA signal, followed by a return to the resting level.

The CTG signals were divided into non-overlapping 20-minute epochs for analysis. Epochs missing more than 20% of samples were not analyzed. Infants without epochs with sufficient data were excluded from the analysis. Fig 2 shows a valid epoch of an FHR trace from a fetus with a healthy outcome [8]. Our methods paper provides a more detailed description of CTG preprocessing and event identification [9].

**Fig 2.**
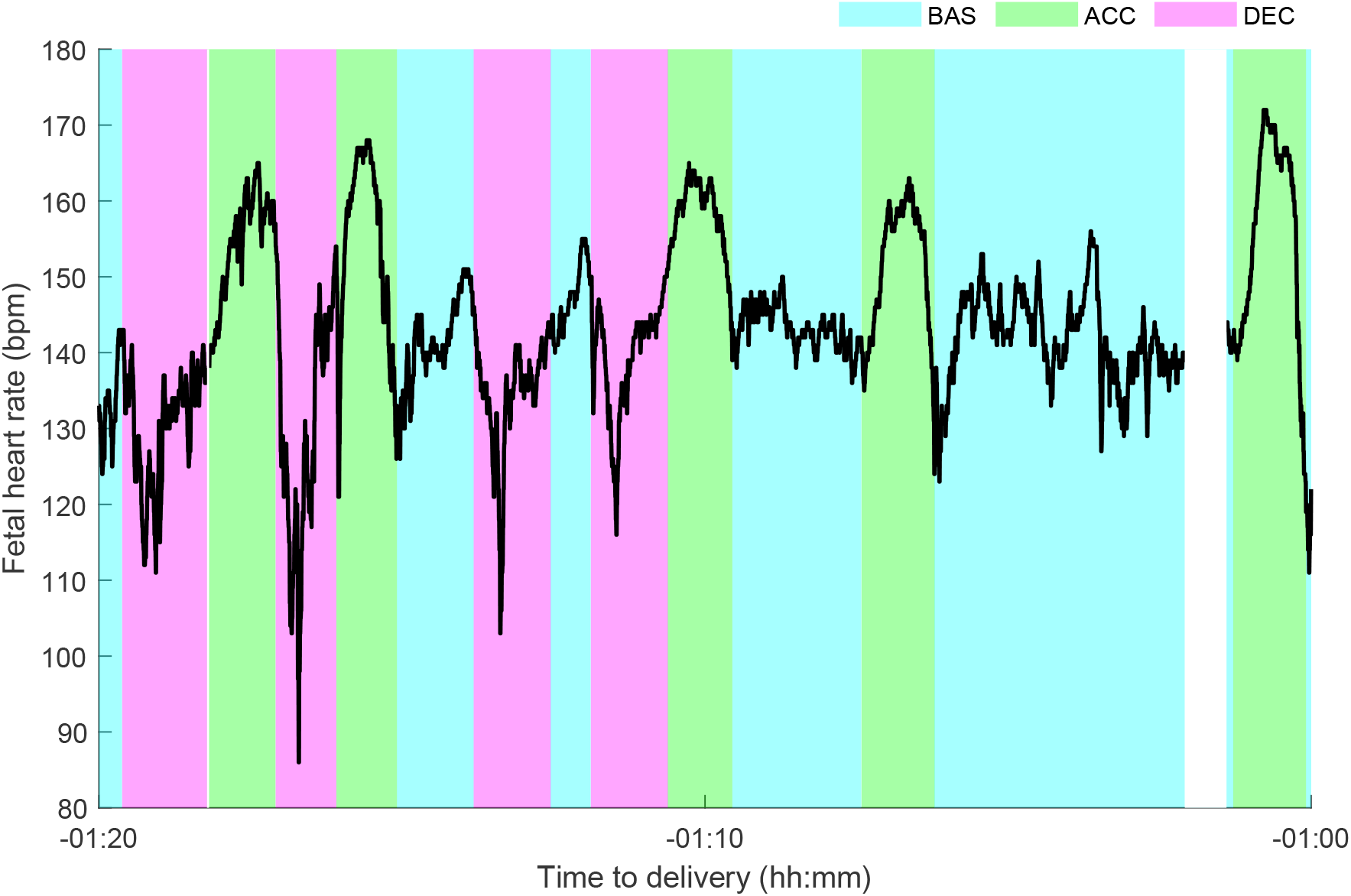
FHR patterns of an epoch starting one hour and 20 minutes before delivery and ending one hour before delivery. The FHR is from a healthy fetus. PeriCALM Patterns identified sections of FHR baseline (cyan), acceleration (green), and deceleration (magenta). Blank spaces correspond to gaps or uninterpretable FHR segments which were removed from analysis.

### Extraction of CTG features

Each FHR epoch was segmented into baseline, acceleration, and deceleration events as labeled by PeriCALM Patterns. We computed FHR and UA features for each event segment. The set of extracted features was based on those that our previous work demonstrated to be associated with fetal outcome [8]. Table 1 lists the CTG features extracted from each event type. The indices indicated in the table will be used in the results section to represent each feature. We extracted a total of 40 features from each CTG epoch [9] from the following categories: event descriptive, time-domain, frequency-domain and nonlinear.

**Table 1.**
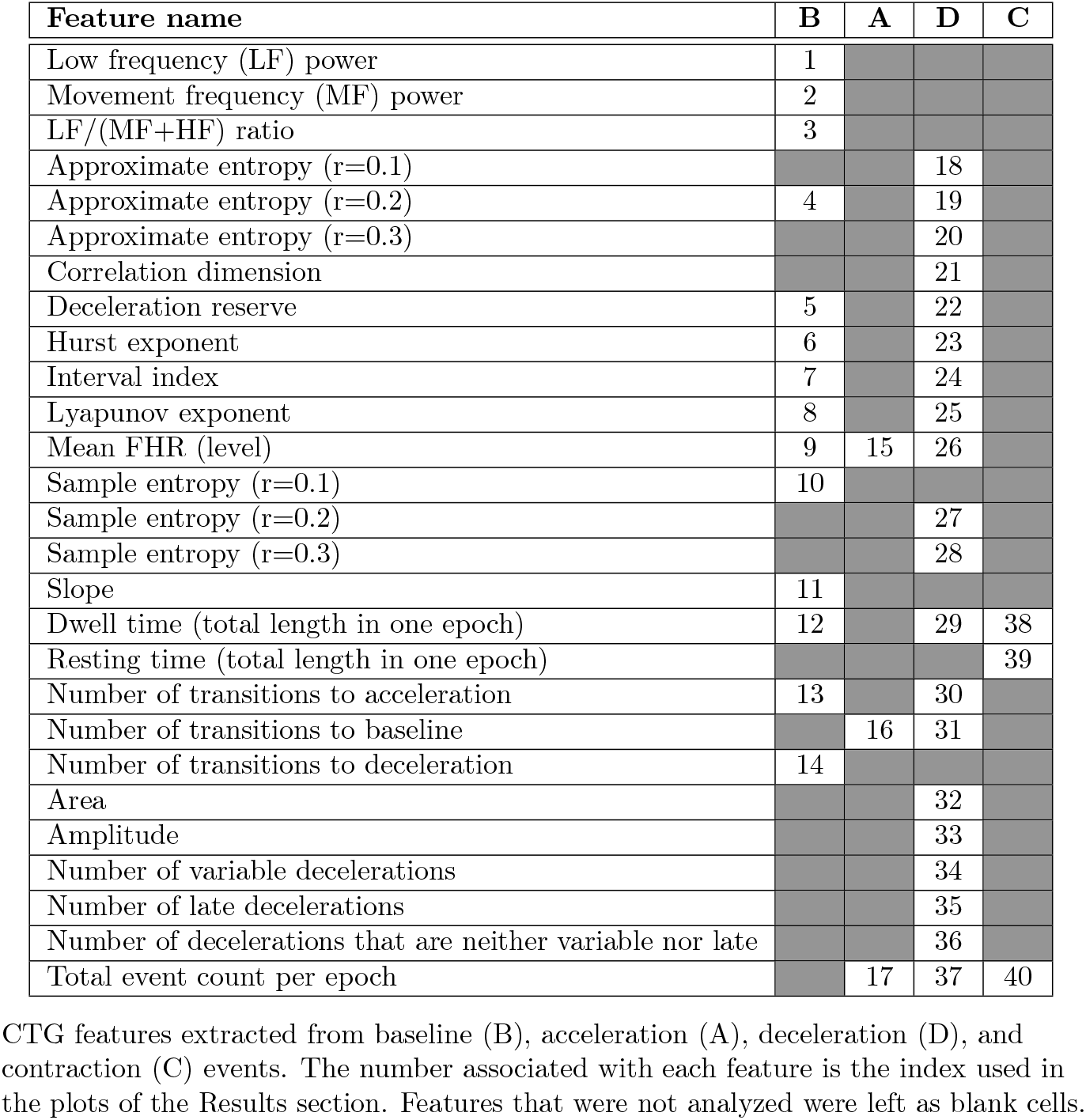
List of CTG Features Extracted.

| Feature name | B | A | D | C |
| --- | --- | --- | --- | --- |
| Low frequency (LF) power | 1 |  |  |  |
| Movement frequency (MF) power | 2 |  |  |  |
| LF/(MF+HF) ratio | 3 |  |  |  |
| Approximate entropy (r=0.1) |  |  | 18 |  |
| Approximate entropy (r=0.2) | 4 |  | 19 |  |
| Approximate entropy (r=0.3) |  |  | 20 |  |
| Correlation dimension |  |  | 21 |  |
| Deceleration reserve | 5 |  | 22 |  |
| Hurst exponent | 6 |  | 23 |  |
| Interval index | 7 |  | 24 |  |
| Lyapunov exponent | 8 |  | 25 |  |
| Mean FHR (level) | 9 | 15 | 26 |  |
| Sample entropy (r=0.1) | 10 |  |  |  |
| Sample entropy (r=0.2) |  |  | 27 |  |
| Sample entropy (r=0.3) |  |  | 28 |  |
| Slope | 11 |  |  |  |
| Dwell time (total length in one epoch) | 12 |  | 29 | 38 |
| Resting time (total length in one epoch) |  |  |  | 39 |
| Number of transitions to acceleration | 13 |  | 30 |  |
| Number of transitions to baseline |  | 16 | 31 |  |
| Number of transitions to deceleration | 14 |  |  |  |
| Area |  |  | 32 |  |
| Amplitude |  |  | 33 |  |
| Number of variable decelerations |  |  | 34 |  |
| Number of late decelerations |  |  | 35 |  |
| Number of decelerations that are neither variable nor late |  |  | 36 |  |
| Total event count per epoch |  | 17 | 37 | 40 |

CTG features extracted from baseline (B), acceleration (A), deceleration (D), and contraction (C) events. The number associated with each feature is the index used in the plots of the Results section. Features that were not analyzed were left as blank cells.

#### Event descriptive features

These features quantify CTG event attributes in each epoch:

- The <u>dwell time</u> measures the duration of all events of the same type within an epoch. We computed the dwell time for baseline, acceleration, deceleration, and contraction events.
- The <u>resting time</u> measures the total time between uterine contractions for an epoch, and it was specific to the UA signal.
- The <u>number of transitions</u> is the count of the transitions between pairs of FHR events within each epoch.
- The <u>number of each event</u> per epoch.
- The <u>deceleration type count</u> is the count of the decelerations of each type per epoch. Decelerations could be late, variable, or neither [5].
- The <u>area</u> of the FHR curve during all deceleration or acceleration events within each epoch.
- The maximum <u>amplitude</u> of all FHR decelerations or accelerations in an epoch.
- The average <u>slope</u> of all FHR baseline segments in each epoch. This yielded a total of 18 features as described by Table 1.

#### Time-Domain features

A variety of time-domain features have been proposed to assess the FHR [13]. Our previous study demonstrated that only the <u>mean of the FHR</u> signal for each event type was significantly associated with the development of HIE in labor [8]. Thus, we extracted a total of three time-domain features [8].

#### Frequency-domain features

We estimated the FHR power spectrum in baseline segments in three bands: the <u>low-frequency (LF - 30 to 15 mHz)</u> the <u>movement-frequency (MF - 0.15 to 0.5 Hz)</u> band, and the <u>high-frequency (HF - 0.5 to 1 Hz)</u> band [14, 15]. The power was normalized with respect to the signal variance. We also computed the LF/(MF + HF) ratio. The HF power was not included since we previously showed that it was not associated with the development of HIE [8], and it tends to lose any discriminative information in the FHR estimation process from CTG signals [16]. Hence, three frequency-domain features of LF, MF, and the LF/(MF + HF) ratio were included in the analysis.

#### Nonlinear features

Finally, we extracted a set of nonlinear FHR features as proposed in previous studies [17] and found to be associated with fetal outcome [8]. These include:

- The <u>approximate entropy (ApEn)</u> compares segments of a signal with itself to measure signal regularity and quantify its randomness [18, 19]. ApEn depends on two parameters: the similarity criterion *r* and embedding dimension *m*. We computed ApEn for *r* = [0.1, 0.2, 0.3] and *m* = 2.
- The <u>sample entropy (SampEn)</u> is similar to ApEn but differs in that its calculation does not depend on the signal length. This makes SampEn robust when comparing signals with different number of samples [20]. SampEn was computed for *r* = [0.1, 0.2, 0.3] and *m* = 2.
- The <u>correlation dimension</u> measures signal regularity, by quantifying signal fractality. This implies finding patterns in the signal that repeat at various scales [17, 21].
- The <u>Lyapunov exponent</u> measures signal regularity by quantifying the dependence of a signal trajectory on its initial conditions [22].
- The <u>Hurst exponent</u> is a measure of signal fractality with a particular focus on long-term behavior [23].
- Phase rectified signal averaging (PRSA) searches for increasing and decreasing tendencies in the signal [24], and aligns and averages all the segments with similar tendencies. The <u>acceleration capacity (AC)</u> is based on the area under the curve of segments with an increasing tendency. In contrast, <u>the deceleration capacity (DC)</u> is obtained from the area under the curve resulting from averaging segments with a decreasing tendency. Finally, the <u>deceleration reserve (DC)</u> is calculated as *DR* = |*DC* | −*AC* and quantifies the dominant tendency of the signal [24].

Sixteen nonlinear features were included in this subset (Table <u>1).</u>

#### Time surrogate variables

We considered four potential surrogate time variables and evaluated their utility:

1. Time to Delivery (TTD): served as a reference variable for comparing the alternative TSVs. TTD quantifies the proximity of the CTG signals to birth. At the onset of labor, the TTD also carries information about the total length of labor.
2. Time from Labor Onset (TLO): aligns signals with respect to the beginning of labor. Thus, it describes how CTG features evolve with the length of labor at any time before delivery. At the time of delivery, the TLO carries information about the total length of labor.
3. Cumulative Contraction Time (CCT): is the total length of contractions that a fetus has endured since the onset of labor. Indirectly, this variable tracks the length of the hypoxic events that the fetus endured, and it serves to assess how the CTG features evolve as the total length of hypoxic events increase.
4. Cumulative Deceleration Time (CDT): tracks the cumulative length of FHR decelerations. Fetal decelerations are the product of reflex responses to strong, long contractions [6]. However, not all contractions are severe enough to cause a fetal response. Thus, the CDT quantifies the length of the events that were severe enough to trigger a fetal reflex response.
5. Contraction Rate (CR): is the number of contractions per epoch. This is a discrete variable that only depends on the current epoch without considering history.

These TSVs were selected because they quantify the progress of labor. The four possible TSVs are independent of delivery method, and can be applied prospectively. In this study, we assessed to what degree each of these TSVs possessed the two other desirable properties mentioned previously: being associated with CTG and improving the association between CTG features and fetal outcome.

#### Mutual information

Mutual information is a measure of linear and non-linear associations among variables [25]. In this study, we used the bivariate and the conditional mutual information to quantify associations among the CTG features ***F***_*i*_, for 1 ≤ *i* ≤ 40, the time surrogate variables ***T***_*j*_, for 1 ≤ *j* ≤ 5, and the fetal outcome ***C*** (healthy without blood gas, healthy with blood gas, acidosis, or HIE). The value of the mutual information is limited by the entropy of the variables. The entropy of a variable is a measure of its uncertainty; it is also understood as its information content. For a variable that follows a Gaussian distribution, its entropy is directly proportional to the log variance. Thus, the entropy of a variable is related to its variability and can be used to normalize the mutual information between 0 and 1 [25, 26].

#### Temporal dependence of the CTG features

To assess the dependence of the CTG features on the TSVs, we computed the mutual information between the features and the time variables, normalized by the entropy of the features. First, we ensured that each feature ***F***_*i*_ was discrete with values *f*_*i*_, as described in the discretization section below. Then we computed the entropy as

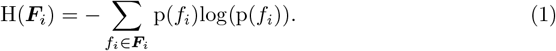

Following this, we computed the mutual information between each feature ***F***_*i*_ and TSV ***T***_*j*_ over all *f*_*i*_ and time steps *t*_*j*_ as

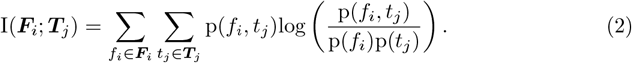

Finally, by defining NMI as the normalization of this mutual information by the feature entropy, it can be shown that the following holds:

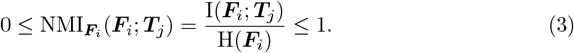

Thus, the 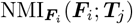 quantifies the proportion of the entropy of each feature is explained by each TSV.

#### Associations between CTG features and fetal outcome

The associations between individual CTG features and the fetal outcome were quantified as NMI_***C***_(***C***; ***F***_*i*_), using the same notation as Eq 3. This NMI quantified the proportion of the entropy of the outcome groups that could be explained by a CTG feature alone. Thus, it quantifies how well a feature could predict fetal outcome [27].

#### Conditional mutual information

The conditional mutual information between two variables given a third one quantifies the added advantage of accounting for that third variable when studying the association between the first two [26]. We computed the conditional mutual information

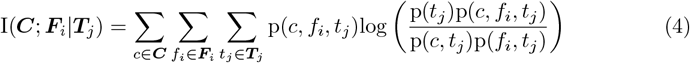

to quantify the gain in information from accounting for the TSV when studying the association between fetal outcomes and CTG features.

From Eq (4) it can be shown that when ***C*** and ***F***_*i*_ are independent of the TSV ***T***_*j*_, then I(***C***; ***F***_*i*_|***T***_*j*_) = I(***C***; ***F***_*i*_). That is, accounting for the TSV does not increase the information a feature provides about the fetal outcome. Also, if ***F***_*i*_ is perfectly correlated with ***T***_*j*_, then I(***C***; ***F***_*i*_|***T***_*j*_) = 0 because once ***T***_*j*_ is given, ***F***_*i*_ provides no additional information about the fetal outcome. Finally, if I(***C***; ***F***_*i*_ | ***T***_*j*_) *>* I(***C***; ***F***_*i*_), then accounting for the TSV increases the information a CTG features provide about fetal outcome.

Lastly, the total information that a CTG feature and TSV provide together on the fetal outcome is given by

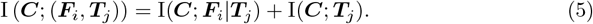

Thus, a candidate TSV would be useful if it is a good indexing variable such that I(***C***; ***F***_*i*_|***T***_*j*_) *>* I(***C***; ***F***_*i*_) and also has a good direct NMI with the fetal outcome I(***C***; ***T***_*j*_).

#### Discretization of continuous variables

Entropy and mutual information are defined for the case of discrete variables in Eq 1, Eq 2, and Eq 4. For continuous variables, they can be approximated from their discretized distribution [25, 26]. Some variables in this study were discrete by definition. For instance, the number of late decelerations per epoch is a discrete CTG feature, and the CR is a discrete TSV. These variables required no further discretization; they were assessed with their original distributions. In contrast, the continuous variables required a careful choice of the number of bins used to discretize their distributions. This required a trade-off between a biased distribution (too few bins) and high random error (too many bins).

We selected the number of bins using Rice University’s discretization rule [28], which depends only on the number of samples and not on the shape of the distribution. Rice’s rule defines the minimum number of bins *N*_bins_ to discretize the distribution as

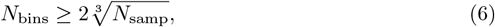

where *N*_samp_ is the number of samples available for a variable [28]. Using this discretization rule ensures that all our features have a similar number of bins, which facilitates cross-feature comparisons. For computing the number of bins, we used the number of epochs in the HIE group, since this was the group with the fewest samples. There were a total of 7,024 HIE epochs, which resulted in *N*_bins_ ≥ 38.3. We used the next power of two, i.e. *N*_bins_ = 64, to discretize the CTG features and TSVs.

Using *N*_bins_, we defined the feature-dependent bin width *w* as

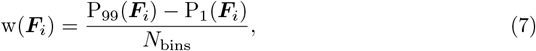

where P_*n*_(***F***_*i*_) is the *n*th percentile of the CTG feature ***F***_*i*_. The same procedure was repeated to discretize the TSVs. The discretization bin width was defined based on the distance between the first and 99th percentiles, then we used that bin width on the complete distribution. This prevented us from using bins that were too wide for variables that had long tails in their distributions or outliers.

The resulting complete set of discrete features ***F***_*i*_, the TSVs ***T***_*j*_, and fetal outcome category ***C*** were used to estimate the entropy and NMI.

### Bootstrapping

We used bootstrapping to generate the distributions of entropy and NMI measures. Bootstrapping was stratified by hospital to maintain intra-hospital variability. Also, since there was a large imbalance among outcome classes, we implemented random subsampling of the larger groups, to prevent the NMI from being biased in favor of the larger groups, especially the healthy without blood gas group which had over 800 times more cases than the HIE group. This procedure consisted in sampling the same number of infants from each group as there were in the minority group - HIE. Thus, the sampling procedure was:

1. For each hospital *k* (1 ≤ *k* ≤ 15):
  a. Count the number of HIE cases *N*_*HIE*_(*k*). The HIE group was the minority class for all hospitals.
  b. Sample randomly with replacement *N*_*HIE*_(*k*) infants from each of the four study groups. This provides a balanced subset of 4 *× N*_*HIE*_(*k*) infants.
2. Aggregate sampled infants across all hospitals.
3. Compute the 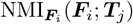, which measures the associations between features and TSVs.
4. Compute NMI_***C***_(***C***; ***F***_*i*_) using the data from all sampled infants. This quantifies the predictability of labor outcomes ***C*** using a stationary analysis of CTG features.
5. Compute the conditional NMI_***C***_(***C***; ***F***_*i*_ | ***T***_*j*_) to assess the usefulness of the TSVs as time indices.

Thus, the NMI measures were computed from a balanced subset of subjects. For the healthy majority group especially, only a small subset was sampled for each bootstrap iteration. However, with a sufficient number of bootstrap iterations almost all infants in all groups were eventually included. We used *N*_*bootstrap*_ = 10, 000 iterations, which have been shown to be sufficient to support statistical comparisons for a significance level *p* < 0.05 [29].

### Statistical tests

The bootstrapping procedure generated the distributions of the NMI statistics for each TSV. These distributions provided measures of the expected value and variability of the NMIs. We used the Friedman test to determine whether the NMI distributions were different among groups. The null hypothesis of the Friedman test is that a set of variables come from the same distribution. We applied this test to the following distributions:

1. H(***F***_*i*_): to compare whether the entropy of the CTG features differed among study groups.
2. 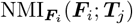: to compare the dependency of CTG features on each TSV, and to determine which TSV was most correlated with the changes in the distribution of CTG features.
3. NMI_***C***_(***C***; ***F***_*i*_ | ***T***_*j*_) and NMI_***C***_(***C***; ***F***_*i*_): to determine whether using a TSV improved the mutual information between an outcome group and feature.

Rejection of the Friedman null hypothesis implied that at least one group of variables was different from the rest. Thus, to confirm the differences among TSVs, we also performed post-hoc pairwise comparisons using the Tukey-Kramer test to protect against family-wise error [30].

## Results

### Study population

Table 2 summarizes the clinical and demographic characteristics of the study groups. Birth weight distributions were similar across groups (*p* = 0.0736). Although statistically significant differences were observed in gestational age, maternal age, and maternal weight, these differences were not large enough to be clinically meaningful. In contrast, nulliparity and length of labor differed substantially among groups. Fetuses who developed HIE were significantly more likely to be born to nulliparous mothers. The ratio of nulliparous to multiparous labors was 4.26 times higher in the HIE group than in the healthy group without blood gas data. Additionally, the median labor duration in the HIE group was 121.2% longer than in the healthy without blood gas group.

**Table 2.** Demographic and Clinical Characteristics of Participants.

|  | Healthy no BG<br>*n* = 129,340 | Healthy with BG<br>*n* = 21,473 | Acidosis<br>*n* = 1,793 | HIE<br>*n* = 155 | *p* |
| --- | --- | --- | --- | --- | --- |
| Birth weight (kg) | 3.39<br>(3.10 - 3.70) | 3.39<br>(3.07 - 3.72) | 3.36<br>(3.07 - 3.69) | 3.43<br>(3.01 - 3.80) | 0.07 |
| Gestational age (weeks) | 39.6<br>(38.9 - 40.3) | 39.7<br>(38.9 - 40.6) | 40.0<br>(39.0 - 40.7) | 40.0<br>(39.1 - 40.7) | $\leq 0.0001$ |
| Maternal age (years) | 31.0<br>(27.1 - 34.4) | 32.0<br>(28.1 - 35.4) | 31.0<br>(26.9 - 34.3) | 31.6<br>(27.4 - 35.2) | $\leq 0.0001$ |
| Maternal weight (kg) | 79.4<br>(70.8 - 90.7) | 78.9<br>(70.3 - 90.3) | 80.7<br>(71.7 - 92.1) | 81.2<br>(72.1 - 96) | $\leq 0.0001$ |
| Nulliparity | 52,056<br>(40.25%) | 11,951<br>(55.66%) | 1,224<br>(68.27%) | 115<br>(74.19%) | $\leq 0.0001$ |
| Length of labor (h) | 8.67<br>(4.87 - 14.20) | 11.33<br>(6.25 - 18.23) | 13.38<br>(8.00 - 21.26) | 19.18<br>(11.02 - 27.33) | $\leq 0.0001$ |

Median (interquartile range) or subject count (percentage) of key characteristics for the four groups. The Kruskal-Wallis test was used on continuous variables and the $\chi^2$ test on categorical data, and *p*-values are reported in the last column.

### Feature entropy

Fig 3 presents the entropy values of individual CTG features across the four study groups. The Friedman test indicated significant differences in the distributions of NMI among groups (*p* ≤ 0.0001). Post-hoc pairwise comparisons confirmed that all groups differed significantly from one another (*p* ≤ 0.0001 for all comparisons). While all comparisons were statistically significant, partly due to the large sample size, Fig 3 shows that the differences among groups were most pronounced for certain feature categories:

**Fig 3.**
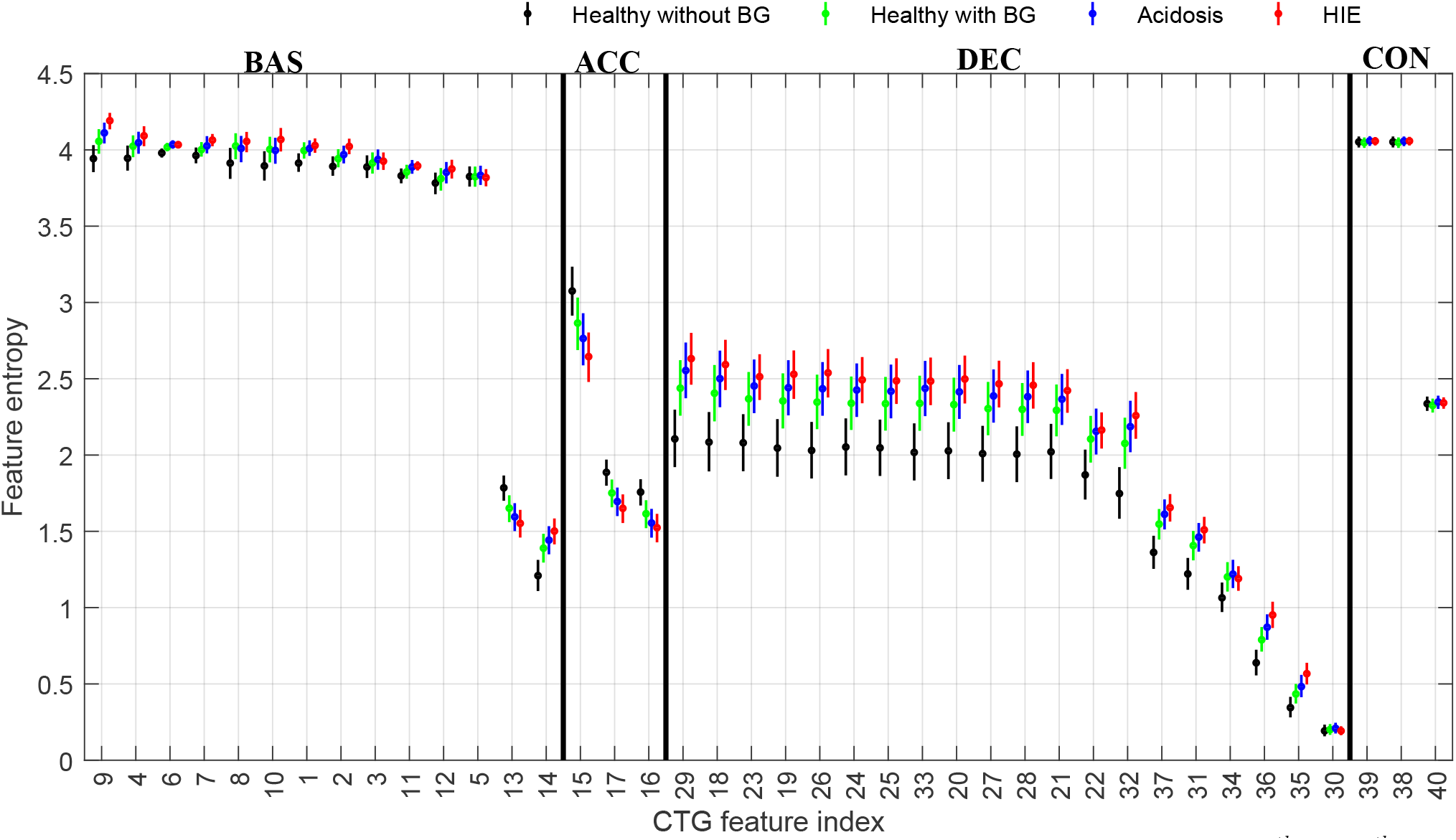
Feature entropy for each event, by outcome group. Range of each feature entropy distribution (2.5^*th*^ − 97.5^*th*^ percentiles, median circled) is shown for baseline (BAS), accelerations (ACC), decelerations (DEC), and contractions (CON) events, and separately for each study group: healthy without blood gas (black), healthy with blood gas (green), acidosis (blue), and HIE (red). The features are sorted in descending order of entropy; indices are listed in Table 1.

1. Baseline feature entropies were highest in the HIE group and lowest in the healthy group without blood gas data, except for feature #13 (number of transitions from baseline to deceleration), which showed the opposite trend.
2. Acceleration feature entropies were highest in the healthy group without blood gas data and lowest in the HIE group.
3. Deceleration feature entropies were highest in the HIE group and lowest in the healthy group without blood gas data.
4. Contraction feature entropies were highest in the acidosis group and lowest in the healthy group without blood gas data.

These findings demonstrate the gradation of the entropy patterns by outcome group severity, particularly for features associated with decelerations and contractions.

### Distribution of time variables

Fig 4 presents the probability density functions of TSV values across all epochs for each study group. The distributions of the TSVs were discretized with Rice’s rule. Fig 4A shows the distribution of TTD revealing that all outcome groups had more observations closer to delivery than earlier in labor. Notably, earlier TTDs were more common in the HIE and acidosis groups compared to the healthy groups. Fig 4B displays the distribution of TLO, with higher values observed more frequently in the HIE and acidosis groups.

**Fig 4.**
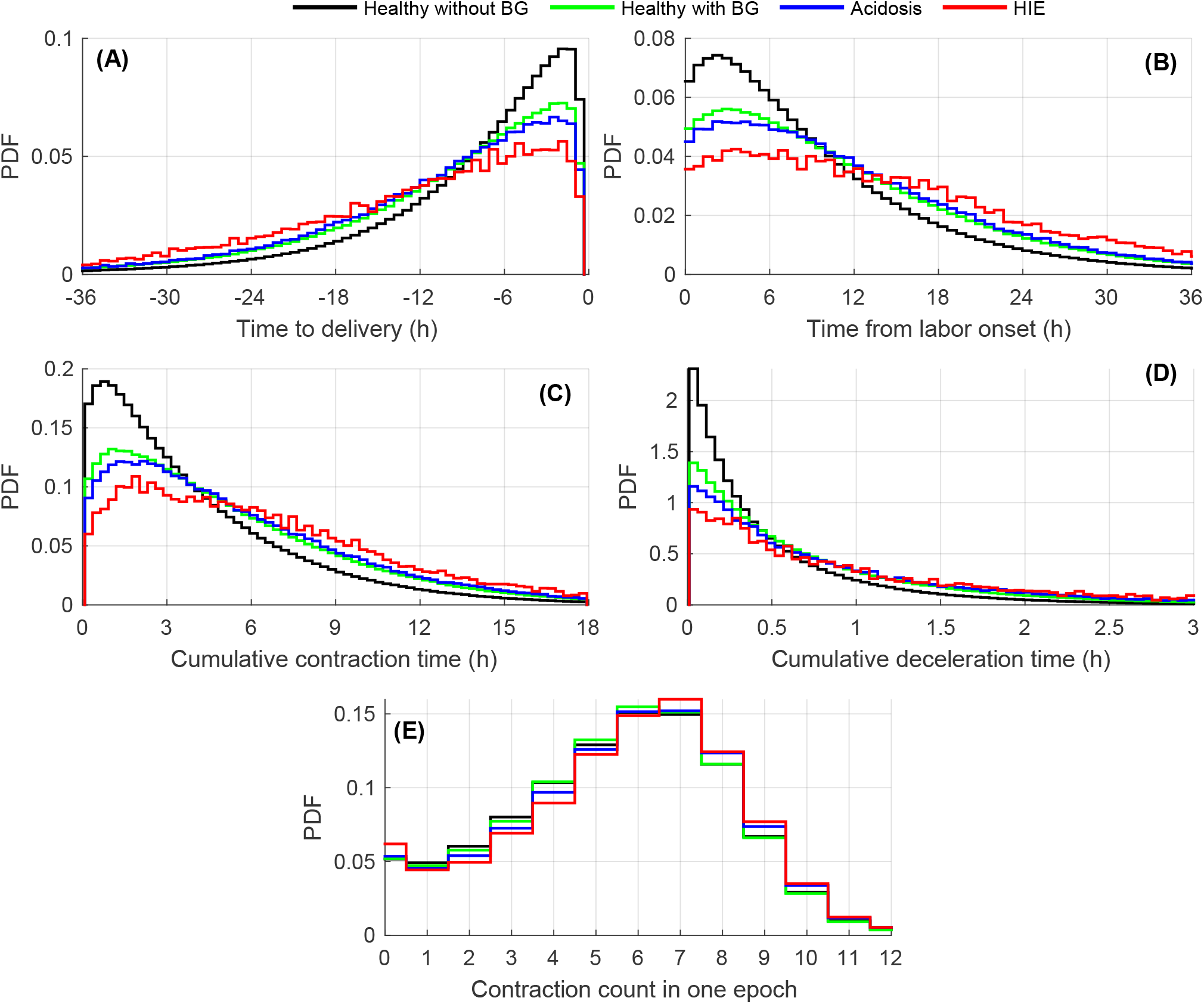
Probability density function of the TSVs. The legend distinguishes outcome groups. (A) Time to delivery (TTD) for last 36 hours before delivery. (B) Time from labor onset (TLO) for first 24 hours of labor. (C) Cumulative contraction time (CCT) up to 18 hours. (D) Cumulative deceleration (CDT) time up to 3 hours.(E) Contraction rate per epoch. We removed the zero values in the CCT and CDT cases, which would mask the rest of the distribution.

Fig 4C and Fig 4D show the distributions of non-zero CCT and CDT, respectively. Both TSVs exhibited similar patterns to TLO, with the HIE and acidosis groups showing longer cumulative durations than the healthy groups. These plots exclude epochs with zero values for CCT and CDT. Specifically, CCT was zero in 5.0% of epochs in the healthy without blood gas group, 4.5% in the healthy with blood gas group, 4.6% in the acidosis group, and 4.7% in the HIE group. CDT was zero in 62.1% of epochs in the healthy without blood gas group, 56.2% in the healthy with blood gas group, 54.1% in the acidosis group, and 52.0% in the HIE group.

Finally, Fig 4E illustrates the distribution of CR per epoch. While the distributions were generally similar across groups, there was a slight trend toward higher CR in the HIE and acidosis groups compared to the healthy groups.

Multivariate Kruskal-Wallis tests rejected the hypothesis of similarity among groups for the distribution of each TSV (*p* ≤ 0.0001 for all cases). Thus, each TSV carried some information about fetal outcome.

### Association between CTG features and TSVs

CTG features had varying degrees of associations with the different TSVs. As Fig 5 shows, deceleration features were most associated with the CDT; their 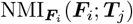 was notably higher than with any other TSV. Contraction features had the highest NMI with the CR but had the lowest NMI for all other features. Feature #40, total number of contractions per epoch is not shown since it coincides with the definition of CR and has 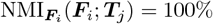. The remaining baseline and acceleration features had similar overlapping NMIs for all other TSVs.

**Fig 5.**
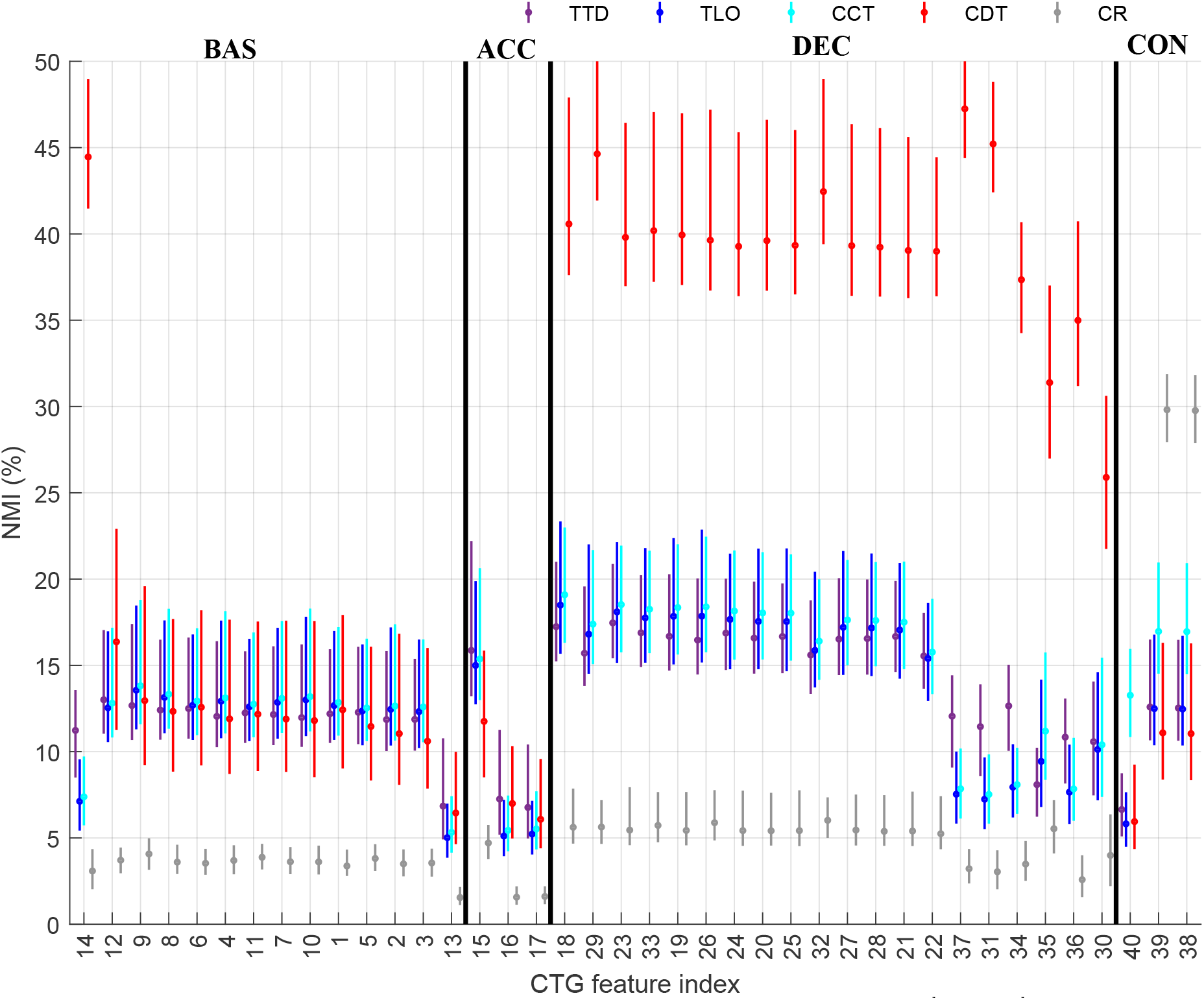
Association between CTG features and TSVs. Range of NMI distributions (2.5^*th*^ − 97.5^*th*^ percentiles, median circled) between the CTG features and each TSV (indicated in legend): the time to delivery (TTD), the time from labor onset (TLO), the cumulative contraction time (CCT), the cumulative deceleration time (CDT), and the contraction rate (CR). The NMI ranges are separated for baseline (BAS), acceleration (ACC), deceleration (DEC), and contraction (CON) events. Features sorted in descending order of NMI; indices listed in Table 1.

When assessing the differences among TSVs for each CTG segment from which the features were calculated, the Friedman test and post-hoc comparisons revealed that all groups were significantly different (*p* ≤ 0.0001). When looking at the systematic behavior of the TSVs across all features, Fig 5 shows that the TLO, the TTD, CCT, and CDT had similar and overlapping 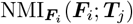 distributions for most features. Thus, all these TSVs were significantly associated with the evolution that CTG features undergo during labor.

### Advantage of accounting for the TSVs

Fig 6 compares NMI_***C***_(***C***; ***F***_*i*_ | ***T***_*j*_), the added advantage of accounting for each TSV when using CTG features to distinguish the outcome groups. There are very clear differences among the TSVs.

**Fig 6.**
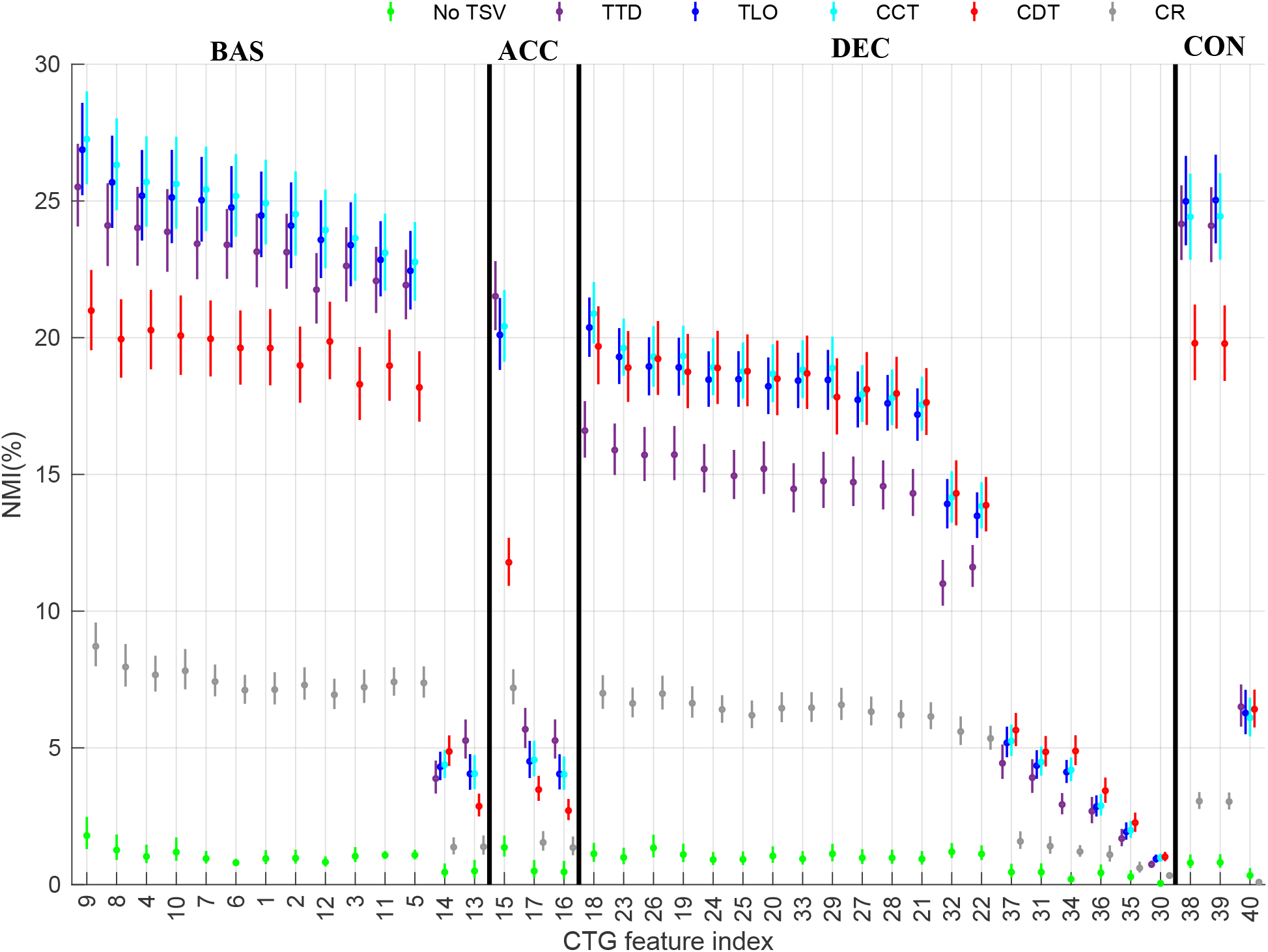
Information provided by CTG features and TSVs on fetal outcome. Range of NMI distributions (2.5^*th*^ − 97.5^*th*^ percentiles, median circled) between CTG features and fetal outcomes for TSV strategies or no TSV (green), time to delivery (TTD, purple), time from labor onset (TLO, blue), cumulative contraction time (CCT, cyan), cumulative deceleration time (CDT, red), and contraction rate per epoch (CR, gray); and grouped by event: baseline (BAS), acceleration (ACC), deceleration (DEC), and contraction (CON). Features sorted in descending order of NMI; indices listed in Table 1.

First, it is clear that use of any of the five TSVs improved NMI, compared to not using a TSV (green symbols). Secondly, the conditional NMI for contraction rate was the lowest of the five TSVs and would be of little use in classification systems. The NMI for TLO and the CCT always overlapped and were the highest for almost all features. The remaining TSVs - TTD and CDT - were among the top TSVs for less features than the previous two. The CDT yielded lower NMIs than the CCT, TLO, and TTD in baseline-, acceleration, and contraction-originated features but performed well for DEC features. In contrast, the TTD yielded lower NMIs than the CCT, CDT, and TLO in deceleration-originated features.

The Friedman test confirmed that there were statistical differences among the groups and that all pairwise differences were also significant with *p* ≤ 0.0001. Thus, when considering the performance across all features, the TLO and the CCT performed best.

## Discussion

This study evaluated five prospective time-surrogate variables (TSV) to determine the best CTG-indexing method for the intrapartum assessment of the risk of HIE. This analysis was performed on data from 152,761 births, one of the largest cohorts of intrapartum data to date. The best TSV should most enhance the associations between CTG features and the outcomes of labor, and for prospective use should be readily available during labor. Our main results are two: (1) using any TSV is better than using none, and (2) the best TSVs for prospective use were the TLO and the CCT. However, TLO is the most practical TSV; tracking it only requires the identification of the onset of labor, which makes it suitable for real-world applications. Therefore, future automated studies aiming for early detection of intrapartum HIE should account for TLO. Doing so has the potential to improve their performance by informing the classifier about the dependence of the discriminability of CTG features on the progress of labor.

### Importance of the time from labor onset in intrapartum CTG assessment

In a previous study, we demonstrated that the association between CTG features and the development of HIE was strongly enhanced by accounting for the TTD during the last six hours of labor [8]. Despite this advantage, TTD has limited applicability: it cannot be used prospectively because the time of delivery is only known after birth. Also, TTD does not reflect the stress of labor. The length of labor varies widely across different births. For example, for labors of one-hour and 12-hour durations, the last hour of labor is associated with different levels of cumulative stress. Finally, TTD does not have the same meaning for vaginal and Caesarean deliveries. In vaginal deliveries, TTD represents the time before the natural conclusion of the progress of labor. In contrast, the time of birth in Caesarean deliveries is determined by a clinical decision; TTD is not associated to the progress of labor in these cases.

In this study, as alternatives to TTD, we examined the use of TLO, CCT, CDT, and CR. Furthermore, we expanded the analysis window and included up to 72 hours of data before delivery. Unlike TTD, none of the proposed TSVs depend on the time of birth, so all can be used prospectively. TLO, CCT, and CDT begin at zero when labor commences; and the contraction rate only depends on the contractions observed within each epoch. TLO tracking requires only the identification of labor onset. In contrast, the other TSVs require the correct identification of contraction or deceleration events, counting the number of events per epoch, and summing the total duration of these event throughout labor. While possible, such procedures are cumbersome for clinical use without specialized software tools. Thus, regardless of the numerical results, we propose that TLO is the most practical TSV to implement in prospective studies.

Our numerical results determined that the TLO and the CCT were the TSVs that most increased the associations between CTG features and fetal outcomes. They were also among the top performers in the assessment of the associations between CTG features and TSVs. Moreover, the differences between these two TSVs were small: they had overlapping distributions for almost all comparisons. Therefore, when taking into account all criteria for a good TSV, we conclude that TLO is the best of those we tested. TLO is available prospectively, tracking it only requires the detection of labor onset, it had significant associations with the CTG features throughout labor, and it largely increased the associations between CTG features and fetal outcomes.

### Implications for classification studies

FHR and UA are inherently nonstationary signals their features exhibit meaningful variation throughout the course of labor [8, 31, 32]. Despite this, classification studies often treat these signals as stationary. Most classifiers are trained on data from the final minutes before delivery, when signs of severe hypoxemia in the FHR are most pronounced [33, 34]. While such models may perform well near the end of labor, their predictive accuracy cannot be reliably extended to earlier time intervals due to the nonstationary nature of CTG. Some studies have attempted to broaden the analysis window to include the end of the first and second stages of labor [35, 36], or the initial moments of CTG monitoring [37]. However, these efforts have not validated performance across the full duration of labor. This lack of comprehensive validation limits the real-world applicability of such systems. A key barrier to broader validation has been the absence of a suitable time variable that accounts for the evolving nature of CTG signals.

Accurate discrimination of nonstationary signals such as FHR and UA requires time-varying models. As shown in Fig 6, incorporating any of the TSVs studied significantly increased the association between CTG features and labor outcomes compared to models that excluded temporal context. As discussed above, TLO was the best and most practical TSV of the set. TLO also aligns with standard clinical practice, where longer labors typically prompt closer monitoring and stricter evaluation of risk factors.

Given its availability throughout labor and its strong association with CTG features and outcomes, we recommend that future classifiers incorporate TLO when analyzing CTG signals. This could be achieved by:

1. Inclusion as a classification feature: TLO could be included explicitly as a classification feature for machine learning classifiers. Thus, these models would learn from the joint distributions and interactions between CTG features and TLO. This approach is the most straightforward to implement in current models.
2. Direct modulation of classification parameters: TLO could be used to dynamically adjust the parameters of a classification model. For instance, in a neural network, the TLO could be presented at each layer of the network. Thus, the weights of each layer would be modulated by the TLO. This way, the classifier would adjust its interpretation of the CTG signals as labor progresses.
3. Temporal stratification of classification models: in this approach, TLO could be discretized, requiring separate classification models for each discrete TLO value, e.g., every 2 hours. This way, the classifiers will learn the differences among groups at different times in labor, and these could be used sequentially for new observations. The strength of this approach would lie in its interpretability, clinical intuition that risk evolves over time would be represented by classifiers that are different for different times.

Although we consider that TLO to be the best TSV for real-time application, we should not disregard the CCT and CDT. While cumbersome to track, the CCT was also a top performer among the TSVs. Future fetal monitoring systems could be designed to identify and track contractions automatically. Also, the CDT was shown in 5 to modulate the FHR decelerative response during labor, and it could be used as a scheduling variable in the development of time-varying UA-FHR models. The best way to track and utilize these cumulative times in future classification models is yet to be explored. Deep-learning models are constantly increasing in size and they are able to learn better from the underlying patterns in CTG signals. Thus, future models could be

pretrained to recognize and quantify FHR decelerations and UA contractions. Then, a classification head could be appended to this event-detection model, drawing from the event information to better interpret the changes in the FHR, UA, and their interaction.

### Outcome-dependent entropy of CTG features

The entropy of a variable measures its uncertainty, and it is proportional to its variability. Fig 3 showed that most CTG features had a similar entropy for each segment from which they were estimated: baseline, acceleration, deceleration, or contractions. Features estimated from baseline segments had the highest entropy. The entropy of features from baseline and contraction events were quite similar across classes. In contrast, the entropy of features from acceleration events were higher in the healthy groups and lowest in the acidosis and HIE groups. Accelerations are markers of a healthy fetal status [5]. We found that they were more prevalent in the healthy group, which could explain their increased entropy. Similarly, features from deceleration events had a higher entropy in the HIE and acidosis groups than the healthy groups. FHR decelerations signal transient fetal hypoxia, and are more common in those fetuses developing acidosis and HIE [32]. Thus, it is sensible that these groups had deceleration features with higher entropies. These results agree with the clinical interpretation that accelerations are most useful in the assessment of healthy cases, and decelerations are most useful in the assessment of pathological cases.

Furthermore, Fig 5 displays an interesting finding: the entropy of all deceleration-originated features was better explained by the CDT than by other TSVs. While the CDT quantifies the progressive length of decelerations endured up to the beginning of the current FHR epoch, the deceleration-based features describe the FHR properties within such epochs. Thus, the relationship between deceleration features and the CDT could suggest the presence of memory where the response to hypoxemia is modulated by past responses. Past animal studies have shown that the properties of FHR decelerations vary due to fetal exposure to repeated hypoxic events [38, 39]. Our results suggest that this is also the case in humans. FHR decelerative response may be modulated by past exposure to deceleration-triggering stimuli. In the past, system identification studies have attempted to model FHR decelerations from uterine contractions [40, 41]. Thus, the CDT could prove to be an important modulator of the parameters of such systems. In summary, accounting for time is necessary during intrapartum monitoring. Failure to do so disregards crucial properties of these signals that enhance their discriminability for fetal hypoxemia and risk of developing HIE.

## Limitations and future work

While our study demonstrated the benefits that TSVs bring, it is important to acknowledge key limitations. Specifically, the acidosis and HIE groups suffer from noisy outcome labels. Our goal was to examine CTG progression linked to fetal deterioration, but early epochs in these groups may exhibit normal FHR patterns prior to the onset of hypoxic stress. These epochs are still labeled as acidosis or HIE during training, potentially obscuring class differences and reducing the NMI between FHR features and fetal outcomes. Despite this uncertainty, early labor data cannot be excluded. The primary objective of CTG analysis is to identify fetuses at high risk of HIE as early as possible to enable timely clinical intervention. Including the full span of labor, even with imperfect labels, may help uncover early predictive signals of fetal compromise. Thus, while noisy labels may lower the quantitative NMI values, they may still support the identification of robust, early-warning features.

It is also important to clarify that our results do not directly reflect prediction accuracy in CTG classifiers. NMI quantifies feature relevance and association with outcomes, but classification performance depends on the interplay of multiple, potentially correlated features. Therefore, classification studies must first assess feature importance and redundancy to select an optimal subset. Then, by neglecting or accounting for the TLO, they should confirm whether the enhanced information on the outcome of labor translates into improved classification performances.

## Conclusion

We evaluated TSVs using one of the largest intrapartum cohorts studied to date. Our findings showed that incorporating any TSV improved the association between CTG features and labor outcomes compared to using no time variable. Among the four proposed TSVs, TLO was a top performer and is far more practical for prospective use than the others. We therefore conclude that TLO is the most useful TSV for CTG indexing in intrapartum studies. Incorporating TLO into future classification models is likely to enhance early-warning systems for HIE risk, enabling timely clinical interventions when they are most effective.

## Data Availability

The datasets generated and analyzed during the current study are not publicly available due to patient privacy policies. Unidentified cardiotocography (CTG) records may be available for research but require submitting a study proposal. The CTG data cannot leave KPNC firewalls under any circumstances. Interested parties might follow these steps: (1) submit a proposal to KPNC Division of Research that includes the author MK (or his designate) as a co-investigator, (2) the proposal will be reviewed and approved by the KPNC Institutional Review Board for the Protection of Human Subjects, and (3) a Data Use Agreement between the interested parties and the Division of Research will be executed. The interested parties must commit to research use only, with no commercial use of the datasets.

## Acknowledgments

We are grateful for the administrative support provided by: Brigid Acuna, Aditi Lahiri, Jennifer Baker, and Eileen Walsh.

We also want to extend our deep appreciation for the help and feedback provided by Dr. Emily Hamilton in the writing of this article.

